# Pre-exposure Prophylaxis Uptake Concerns in the Democratic Republic of the Congo: Key Population and Healthcare Workers Perspectives

**DOI:** 10.1101/2023.01.13.23284513

**Authors:** Yanhan Shen, Julie Franks, William Reidy, Halli Olsen, Chunhui Wang, Nadine Mushimbele, Richted Tenda Mazala, Tania Tchissambou, Faustin Malele, Apolinaire Kilundu, Trista Bingham, Gaston Djomand, Elie Mukinda, Raimi Ewetola, Elaine J. Abrams, Chloe A. Teasdale

## Abstract

Key populations (KP) in the Democratic Republic of the Congo (DRC), including female sex workers (FSW), are disproportionally affected by HIV. Quantitative feedback surveys were conducted at seven health facilities in DRC with 70 KP clients enrolled in services to measure pre-exposure prophylaxis (PrEP) benefits and concerns. The surveys also assessed satisfaction with PrEP services and experiences of stigma at the health facilities. Thirty healthcare workers (HCW) were surveyed to measure attitudes, beliefs, and acceptability of providing services to KP. KP client survey participants were primarily female SW. KP clients reported that the primary concern about taking PrEP was fear of side effects. HCW concurred with clients that experienced and anticipated side effects were a primary PrEP uptake concern, along with costs of clinic visits.

## INTRODUCTION

Key populations (KP), including sex workers (SW), men who have sex with men (MSM), people who inject drugs (PWID) and transgender (TG) women, are disproportionately affected by HIV (UNAIDS, 2021). In 2019, KP and their sexual partners were estimated to have accounted for 65% of new adult infections worldwide (1). For KP who are HIV-negative, expanding access to and uptake of pre-exposure prophylaxis (PrEP) is a key HIV prevention strategy. PrEP can greatly reduce the incidence of HIV infection (2–6), however expanding access to PrEP in many resource limited settings (RLS) remains challenging (7). While supply and cost issues are important factors (5, 7–11), hesitancy among healthcare workers (HCW) and clients may also be a barrier to PrEP access and uptake (12, 13).

In 2020, the Democratic Republic of the Congo (DRC) had an estimated HIV prevalence of 0.7% (95% CI: 0.6%-0.9%) among adults aged 15 to 49 years (14). KP in DRC, particularly FSW, are disproportionally affected by HIV, having higher rates of new infections (14). In 2020, HIV prevalence among SW was 7.5%, which was nine times higher than in the general population (14–16). We report data from surveys conducted with KP clients accessing PrEP services in health facilities in DRC measuring client and healthcare worker (HCW) perceived benefits and concerns about PrEP, as well as HCW attitudes and acceptability of providing HIV services to KP.

## METHODS

In collaboration with the DRC Ministry of Health and the Centers for Disease Control and Prevention (CDC), ICAP at Columbia University conducted the first pilot of PrEP services in DRC. Results on PrEP uptake from a study have been previously reported (17). The project was conducted between February and November of 2018 at seven ‘KP-friendly’ health facilities in Kinshasa (four facilities) and Lubumbashi (three facilities), which receive support from the CDC through the U.S. President’s Emergency Plan for AIDS Relief (PEPFAR) to provide health services for KP clients. To support the implementation of PrEP, health facilities used a comprehensive training package developed for the pilot, which includes a four-day training curriculum for HCW and clinic staff, monitoring and evaluation tools for clinic-level and national reporting of PrEP services, standard operating procedures (SOPs), and job aids for HCW (18). HCW also received the CDC’s HCW sensitization training to increase their understanding of the unique medical and psychosocial needs of KP clients and were trained to use the PEPFAR *Monitoring, Evaluation, and Reporting (MER) 2*.*0 KP Classification Tool* to help identify KP clients eligible for PrEP (19).

At the end of the pilot project, quantitative feedback surveys were conducted at the seven project health facilities to measure satisfaction and to identify concerns about PrEP services among KP respondents ≥18 years who were able to understand French, Lingala or Swahili. Ten KP clients who had initiated PrEP were sampled per facility using convenience sampling, including clients currently taking PrEP and those who had discontinued. Details of PrEP initiation are described elsewhere (Franks et al., 2021). The survey was interviewer-administered and collected demographic information, including sex at birth, self-identified gender, age, and KP group identification based on the PEPFAR *MER 2*.*0 KP Classification Tool*, as well as risk behaviors and HIV testing history. Current concerns about PrEP use were assessed by collecting respondent’s reaction to a 5-item modified Likert scale adapted from published instruments (20–23). Participants reported current PrEP intake, number of days with missed PrEP doses in past seven days, and the reasons for missing doses selected from a list of items. Data were all self-reported including HIV status (medical records were not used to verify information). All participation was voluntary and participants provided verbal consent. Participants received $10 USD for completing the survey.

In addition to the client survey, HCW at the seven project facilities were also invited to participate in a survey about PrEP services for KP. HCW ≥ 18 years, who were French speaking and had at least 3 months of experience providing HIV-related services to KP at the project facilities were eligible. Convenience samples of 5 HCW per facility were recruited. The HCW survey was adapted from the Health Policy Initiative tool to assess HIV-related stigma and discrimination in health facilities and providers (24) and collected information on participant age and sex. HCW perceptions about clients’ PrEP concerns and HCW acceptability of providing services to KP were assessed by a 5-item modified Likert scale (“strongly agree”, “agree”, “don’t know”, “disagree”, “strongly disagree”). HCW opinions towards “what can be done to improve the service provided to KP in health facilities in the DRC” and “what types of training would you recommend for health professionals” were collected (select all that apply). Attitudes about recommending PrEP were collected by asking “Would you recommend PrEP to a patient, friend, or family member?” with response options “Definitely”, “Probably”, “Maybe”, “Probably not”, “Don’t know”. HCW surveys were self-administered on electronic tablets after instructions were given by the study team. HCW participation was voluntary and verbal consent was taken. No compensation was given to HCW and their personally identifiable information was not recorded.

Descriptive data from the surveys are reported. The project was not designed to present results by KP groups, as such results are presented using descriptive statistics without tests of statistical significance. The study protocol was approved by the DRC Ministry of Public Health’s National Ethics Committee for Health, the Columbia University Irving Medical Center Institutional Review Board. This project was reviewed in accordance with CDC human research protection procedures and was determined to be research, but CDC investigators did not interact with human subjects or have access to identifiable data or specimens for research purposes.

## RESULTS

Seventy participants completed the KP PrEP survey; median age was 31 years [IQR: 28 - 38] and 58 (83%) self-identified as female gender (**Table 1**). Almost all (96%) PrEP survey respondents reported sale of sex as a main source of income, 9 (13%) reported injection drug use, and 66 (94%) said that they were on PrEP at the time of the survey.

**Table 1.** Characteristics of key population clients and healthcare workers at 7 health facilities in the Democratic Republic of the Congo

| PrEP Client Survey (N=70) |  | HCW PrEP Survey (N=30) |  |
| --- | --- | --- | --- |
| Sex at birth, N (%) |  | Sex at birth, N (%) |  |
| Female | 38 (54%) | Female | 20 (67%) |
| Male | 32 (46%) | Male | 10 (33%) |
| Self-identified gender, N (%) |  | Age (years), N (%) |  |
| Female | 58 (83%) | < 25 | 1 (3%) |
| Male | 10 (14%) | 25-34 | 20 (67%) |
| Transgender (Male to Female) | 2 (3%) | 35-44 | 5 (17%) |
|  |  | 45-54 | 4 (13%) |
| KP classification<sup>1</sup>, N (%) |  | Professional title as HCW, N (%) |  |
| SW | 38 (54%) | Deputy Director | 2 (7%) |
| TG/SW | 14 (20%) | Doctor of Medicine | 6 (20%) |
| TG/SW/PWID | 6 (9%) | Nurse/Midwife | 11 (37%) |
| TG/PWID | 2 (3%) | Worker/Social worker | 5 (17%) |
| SW/PWID | 1 (1%) | Community counselor | 1 (3%) |
| SW/MSM | 8 (11%) | Laboratory Technician | 1 (3%) |
| MSM | 1 (1%) | Other<br>Refuse to respond | 2 (7%)<br>2 (7%) |
| Age (years), median [IQR] | 31 [28 – 38] | Years working in current position, median [IQR] | 2.5 [2.0 – 5.0] |
| Selling sex as main income, N (%) |  | Years working in current position, N (%) |  |
| Yes | 67 (96%) | 1-2 | 14 (47%) |
| No | 3 (4%) | 3-5 | 12 (40%) |
|  |  | 5-10 | 2 (7%) |
|  |  | > 10 | 2 (7%) |
| Inject drugs, N (%) |  | Years working with HIV+ patients, N (%) |  |
| Yes | 9 (13%) | < 1 | 3 (10%) |
| No | 61 (87%) | 1-2 | 12 (40%) |
|  |  | 3-5 | 12 (40%) |
|  |  | 5-10 | 1 (3%) |
|  |  | > 10 | 2 (7%) |
| Currently on PrEP, N (%) |  | Years providing HIV services for KP, N (%) |  |
| Yes | 66 (94%) | < 1 | 2 (7%) |
| No | 4 (6%) | 1-2 | 15 (50%) |
|  |  | 3-5 | 12 (40%) |
|  |  | 5-10 | 0 |
|  |  | > 10 | 1 (3%) |
| KP PrEP adherence barriers *, N (%) |  |  |  |
| Forgot | 17 (24%) |  |  |
| Worried about side effects | 12 (17%) |  |  |
| Had side effects | 10 (14%) |  |  |
| Ran out of pills | 7 (10%) |  |  |
| Didn't feel like taking it | 4 (6%) |  |  |
| Felt the pills were not needed | 4 (6%) |  |  |
| Other | 8 (11%) |  |  |
| Refusal to respond | 15 (21%) |  |  |

Abbreviations: MSM: men who have sex with men, SW: sex workers, PWID: people who inject drugs, TG: transgender
\*Non-exclusive answers

A total of 30 of 35 HCW participated in the survey; 20 (67%) were female and most (67%) were 25-34 years of age (**Table 1**). About a third of HCW participants were nurses or midwives and a quarter were doctors. About half of HCW who participated in the surveys had been working with people living with HIV (PLHIV) and KP for less than 2 years.

Among KP clients, almost all (90%) reported that taking PrEP would help them and their partners stay HIV negative and 90% reported that taking PrEP would set a good example for other people (**Figure 1A**). The most frequently reported concern among PrEP clients was possible side effects (67%). In addition, 23% of KP were concerned about experiencing HIV-related stigma and 30% worried about losing social support as a result of taking PrEP. Among HCW, actual experiences of PrEP side effects (47%), fear for PrEP side effects (50%), and costs of getting to the clinic would be barriers to client PrEP uptake (**Figure 1B**).

**Figure 1:**
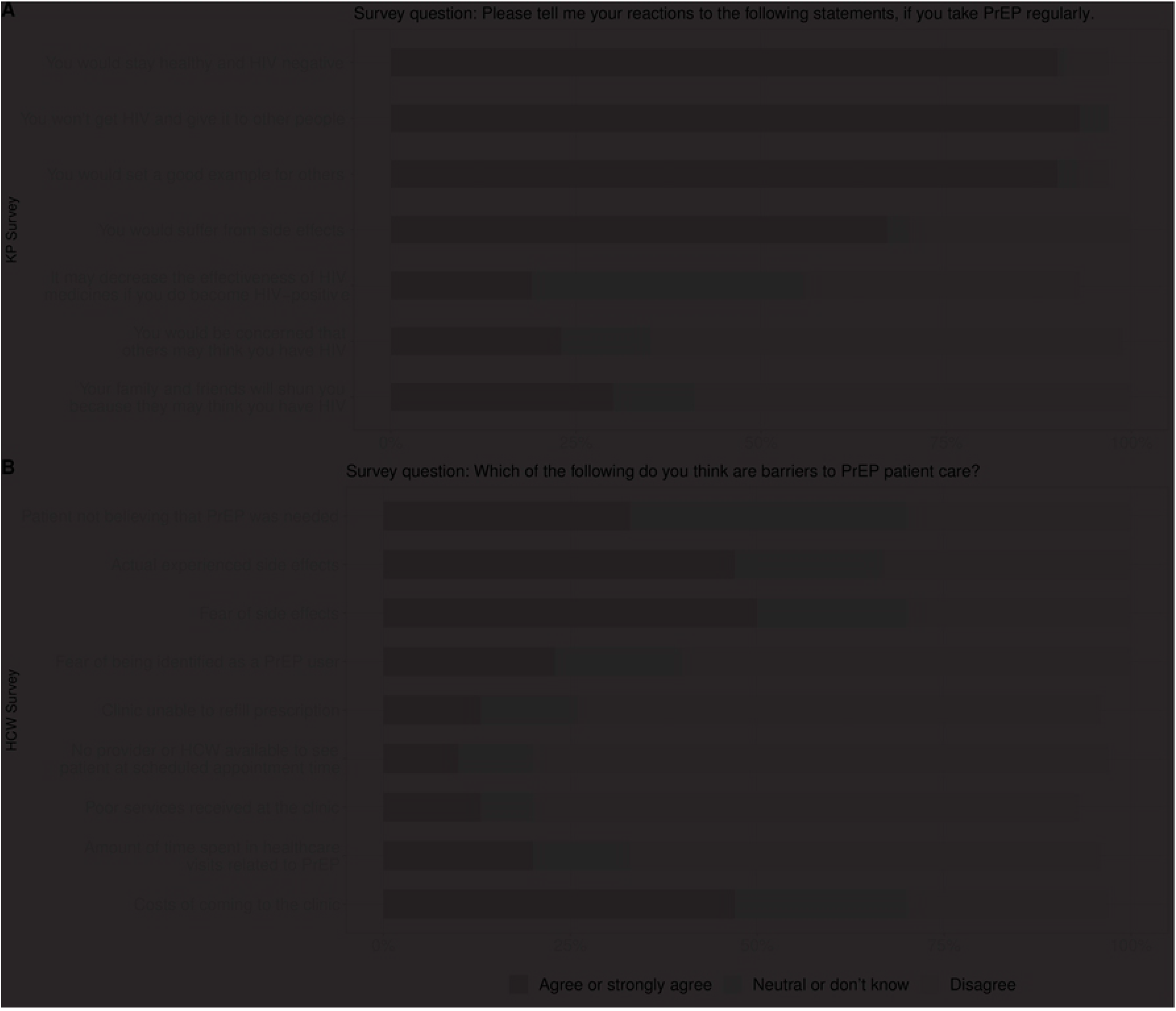
Key population (KP) clients and healthcare workers (HCW) perceived PrEP uptake facilitators and concerns, and KP self-reported PrEP adherence barriers, at 7 health facilities in the Democratic Republic of the Congo Panel A: KP (N=70) perceived PrEP uptake facilitators and concerns Panel B: HCW (N=30) perceived KP PrEP uptake concerns

Reasons for missed PrEP doses over the past 7 days were reported by 55 (79%) of the KP survey respondents (data not shown). The most common reasons for missed doses were forgetting (31%), worrying about side effects (22%), and actually experiencing side effects (18%). In addition, KP also reported running out of pills (13%), not wanting to take (7%), and feeling that PrEP was not needed (7%) as reasons for missing PrEP doses.

All (100%) HCW agreed or strongly agreed that they felt comfortable providing care to SW, TG and MSM, whereas 10% of HCW expressed a neutral attitude and 3% refused to answer towards providing services to PWID (**Figure 2**). Three HCW (10%) reported that KP do not deserve the same quality of healthcare as other patients, and 7 (23%) believed that HIV is a punishment for inappropriate behavior on the part of KP. While the majority (80%) of HCW felt that they were adequately trained to provide high-quality and appropriate care for KP (**Figure 2**), 19 (63%) reported that additional training would help improve services for KP in healthcare facilities (**Supplementary Table 1**). Of note, 23 (77%) and 20 (67%) agreed that clinical competency in providing care for KP and communication with KP could be improved, respectively (**Supplementary Table 1**). Half of HCW said they would definitely recommend PrEP to a patient, and/or friend, and/or family member (data not shown).

**Figure 2:**
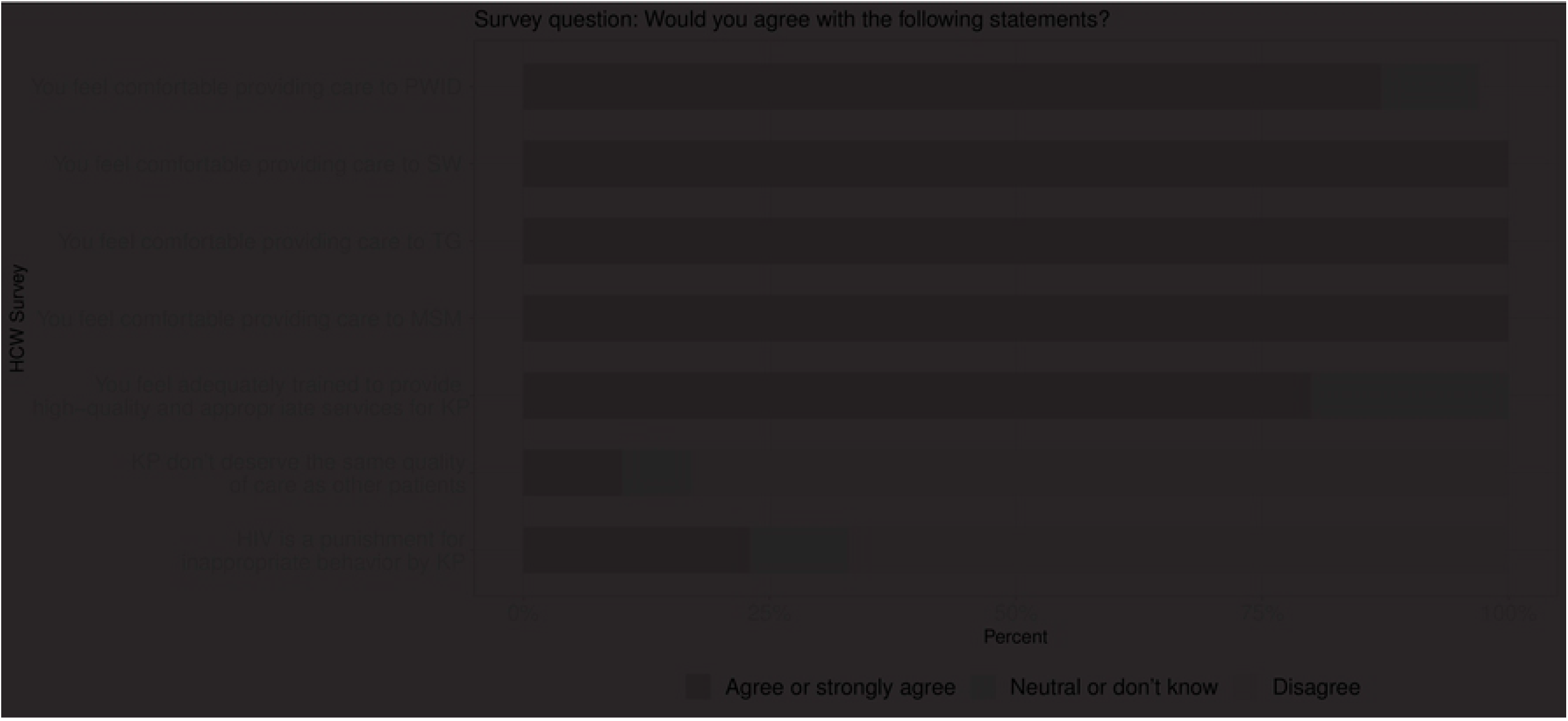
Healthcare workers (HCW) attitudes towards providing services to KP, in the Democratic Republic of the Congo (N=30)

Finally, survey questions about stigma experienced by KP clients showed that 2 (3%) reported having been verbally insulted or harassed within the health facilities, 3 (4%) receiving substandard care, and 1 (1%) reported that they experienced lack of confidentiality during their visit on the day of survey. No KP clients reported being treated disrespectfully by HCW.

## DISCUSSION

In this study of KP, primarily female SW, who were accessing the first available PrEP services in the DRC, most agreed that taking PrEP would protect them and their partners from HIV. In addition, while few KP reported actual side effects as a reason for their own lack of PrEP adherence, many reported fear of side effects and concerns about stigma from family and friends as barriers to taking PrEP. Forgetting to take medication was the most commonly reported adherence barrier and was reported by a third of study respondents taking PrEP. HCW who provide KP health services, including PrEP, were also concerned that perceived and actual experience of side effects would be barriers to client PrEP uptake and adherence. Overall, HCW expressed positive attitudes about PrEP and providing healthcare services to KP.

Similar findings on reasons for and barriers to PrEP uptake have been reported from studies conducted in other African countries. In a qualitative study of female SW and sero-discordant couples in Zimbabwe, perceived HIV risk and concern about acquiring infection were key drivers of PrEP uptake while reasons for declining PrEP included fears of pill burden and side effects, as well as discouragement from family members (25). Qualitative studies in sero-discordant couples in Kenya and female SW in South Africa also found that fear of side effects and stigma were important barriers to PrEP uptake, along with doubts about its effectiveness (26, 27). These findings highlight the importance of educating those who could benefit from PrEP about the low risk of side effects and its overall effectiveness in preventing HIV infection, as well as those who perceive HIV risk and concern about acquiring infection as messengers for PrEP uptake.

Our findings also underscore the continuing challenges related to stigma associated with HIV prevention and treatment interventions, which is further compounded for KP who face additional stigma in this setting. One strategy for improving PrEP uptake and adherence may be to build trust and improve information sharing between HCW and clients (8, 27, 28). While most HCW at these seven health facilities said they would recommend PrEP, they also expressed concerns about side effects and that clients may not believe that PrEP is effective. In a study from Eswatini (formerly Swaziland), HCW reported concerns that PrEP uptake would reduce condom use and cause HIV drug resistance. Providing training for HCW on PrEP is warranted so that they can appropriately counsel clients about the low frequency of side effects and its prevention effectiveness. Also, strengthening virtual or in-person support groups would facilitate sharing personal experience and stories about PrEP effectiveness. Further studies are also needed to assess whether HCW feelings and opinions about PrEP influence client uptake and adherence and what can be done to improve HCW awareness and attitudes.

A strength of our study was collection of data on the views of HCW who provide services to KP. In our survey, all HCW at the seven health facilities reported feeling they had adequate training and felt comfortable providing services to MSM, SW, and TG, however some expressed discomfort with caring for PWID. The HCW in our study work in health facilities that receive support to provide care for KP and received sensitivity training, which may help explain the high acceptability of caring for KP and PrEP. Data from other settings have shown less comfort among HCW for providing services to KP. In South Africa, 30.2%, 25.2%, and 27.7% of HCW strongly felt comfortable providing health services for SW, PWUD, and MSM, respectively (29). Studies from Ghana and South Africa have suggested that stigma-reduction interventions, including training for HCW, can help reduce negative attitudes among HCW towards KP (30, 31). These efforts are critical as client distrust of healthcare providers has been identified as key barriers to PrEP uptake among KP (32, 33). Though there is lack of direct evidence that HCW’s friendly attitude towards KP promote KP PrEP uptake, a study conducted in Kenya showed healthcare providers’ ability to provide high-quality empathetic care have been reported as crucial for improving antiretroviral therapy adherence among MSM living with HIV (34).

There are some limitations of our study, including the relatively small sample sizes of 70 KP and 30 HCW who only were able to understand French, Lingala or Swahili. Furthermore, the convenience samples of participants may have resulted in a sample willing to participate who may have had better experiences at these facilities. In addition, we measured barriers to PrEP adherence among KP clients at one point in time and do not have information about how long those clients had been on PrEP. As noted, the study was also conducted in designated KP-friendly health facilities where staff may be more accepting of KP and supportive of PrEP compared with HCW in clinics serving the general public. As such, our findings may not be generalizable to other settings.

Overall, we found positive attitudes about PrEP among KP clients enrolled in PrEP services and among HCW providing these services. Our results also showed persistent concerns about potential side effects and stigma associated with PrEP. Forgetting to take PrEP was the most commonly reported barrier to adherence, which underscores the need for longer-acting PrEP modalities that are on the horizon. Most HCW at these KP-friendly health facilities reported feeling adequately trained and comfortable providing care to KP clients.

## Data Availability

The study includes data abstracted from medical charts, therefore, the study data belong to the government of DRC so cannot be made publicly available for download. The data can be shared when requested, inquiries should be sent to

## ACKNOWLEDGMENTS

The authors gratefully acknowledge the collaboration of staff members at the KP-friendly clinics in Kinshasa and Lubumbashi whose dedication made this demonstration project possible, as well as the participation of the clinic clients.

This research project has been supported by the President’s Emergency Plan for AIDS Relief (PEPFAR) through the Centers for Disease Control and Prevention (CDC) under the terms of Cooperative Agreement GH000994.

## DECLARATION OF INTEREST STATMENT

No potential conflict of interest was reported by the author(s).

## DISCLAIMER

The findings and conclusions in this manuscript are those of the author(s) and do not necessarily represent the official position of the Centers for Disease Control and Prevention.

## References

1. UNAIDS. New HIV infections among gay men and other men who have sex with men increasing 2020 [Available from: https://www.unaids.org/en/resources/presscentre/featurestories/2020/december/20201207_new-hiv-infections-increasing#:∼:text=people%20who%20injec-,In%202019%2C%20key%20populations%20(including%20gay%20men%20and%20other%20men,other%20than%20eastern%20and%20southern.

2. Sophus AI, Mitchell JW. A Review of Approaches Used to Increase Awareness of Pre-exposure Prophylaxis (PrEP) in the United States. AIDS Behav. 2019;23(7):1749–70.

3. Huang X, Hou J, Song A, Liu X, Yang X, Xu J, et al. Efficacy and Safety of Oral TDF-Based Pre-exposure Prophylaxis for Men Who Have Sex With Men: A Systematic Review and Meta-Analysis. Front Pharmacol. 2018;9:799.

4. Becquet V, Nouaman M, Plazy M, Masumbuko JM, Anoma C, Kouame S, et al. Sexual health needs of female sex workers in Côte d’Ivoire: a mixed-methods study to prepare the future implementation of pre-exposure prophylaxis (PrEP) for HIV prevention. BMJ Open. 2020;10(1):e028508.

5. Bowring AL, Ampt FH, Schwartz S, Stoové MA, Luchters S, Baral S, et al. HIV pre-exposure prophylaxis for female sex workers: ensuring women’s family planning needs are not left behind. J Int AIDS Soc. 2020;23(2):e25442.

6. Lelutiu-Weinberger C, Wilton L, Koblin BA, Hoover DR, Hirshfield S, Chiasson MA, et al. The Role of Social Support in HIV Testing and PrEP Awareness among Young Black Men and Transgender Women Who Have Sex with Men or Transgender Women. J Urban Health. 2020;97(5):715–27.

7. Elliott T, Sanders EJ, Doherty M, Ndung’u T, Cohen M, Patel P, et al. Challenges of HIV diagnosis and management in the context of pre-exposure prophylaxis (PrEP), post-exposure prophylaxis (PEP), test and start and acute HIV infection: a scoping review. Journal of the International AIDS Society. 2019;22(12):e25419–e.

8. Busza J, Phillips AN, Mushati P, Chiyaka T, Magutshwa S, Musemburi S, et al. Understanding early uptake of PrEP by female sex workers in Zimbabwe. AIDS Care. 2020:1–7.

9. Ahouada C, Diabaté S, Mondor M, Hessou S, Guédou FA, Béhanzin L, et al. Acceptability of pre-exposure prophylaxis for HIV prevention: facilitators, barriers and impact on sexual risk behaviors among men who have sex with men in Benin. BMC Public Health. 2020;20(1):1267.

10. Footer KHA, Lim S, Rael CT, Greene GJ, Carballa-Diéguez A, Giguere R, et al. Exploring new and existing PrEP modalities among female sex workers and women who inject drugs in a U.S. city. AIDS care. 2019;31(10):1207–13.

11. Annequin M, Villes V, Delabre RM, Alain T, Morel S, Michels D, et al. Are PrEP services in France reaching all those exposed to HIV who want to take PrEP? MSM respondents who are eligible but not using PrEP (EMIS 2017). AIDS Care. 2020;32(sup2):47–56.

12. Ngure K, Kimemia G, Dew K, Njuguna N, Mugo N, Celum C, et al. Delivering safer conception services to HIV serodiscordant couples in Kenya: perspectives from healthcare providers and HIV serodiscordant couples. J Int AIDS Soc. 2017;20(Suppl 1):21309.

13. Lazarou M, Fitzgerald L, Warner M, Downing S, Williams OD, Gilks CF, et al. Australian interdisciplinary healthcare providers’ perspectives on the effects of broader pre-exposure prophylaxis (PrEP) access on uptake and service delivery: a qualitative study. Sex Health. 2020;17(6):485–92.

14. UNAIDS. Democratic Republic of the Congo HIV and AIDS estimates 2020 2021 [Available from: https://www.unaids.org/en/regionscountries/countries/democraticrepublicofthecongo#:∼:text=Democratic%20Republic%20of%20the%20Congo%20%7C%20UNAIDS&text=In%20the%20Democratic%20Republic%20of,of%20all%20ages%20was%200.21.

15. Kakisingi C, Muteba M, Mukuku O, Kyabu V, Ngwej K, Kajimb P, et al. Prevalence and characteristics of HIV infection among female sex workers in Lubumbashi, Democratic Republic of Congo. Pan Afr Med J. 2020;36:280.

16. Pinto RM, Berringer KR, Melendez R, Mmeje O. Improving PrEP Implementation Through Multilevel Interventions: A Synthesis of the Literature. AIDS Behav. 2018;22(11):3681–91.

17. Franks J, Teasdale C, Olsen H, Wang C, Mushimebele N, Tenda Mazala R, et al. PrEP for key populations: results from the first PrEP demonstration project in the Democratic Republic of the Congo. AIDS Care. 2021:1–4.

18. ICAP. ICAP Pre-exposure Prophylaxis (PrEP) package (Includes training material in French) 2017 [Available from: https://icap.columbia.edu/tools_resources/icap-pre-exposure-prophylaxis-prep-package-2/.

19. PEPFAR. Monitoring, Evaluation, and Reporting (MER 2.0) Indicator Reference Guide 2017 [Available from: https://srhrindex.srhrforall.org/uploads/2018/11/2017_PEPFAR-Monitoring-Evaluation-and-Reporting-Indicator-Reference-Guide_MER-2.0-Version-2.1.pdf.

20. Kalichman SC, Catz S, Ramachandran B. Barriers to HIV/AIDS treatment and treatment adherence among African-American adults with disadvantaged education. J Natl Med Assoc. 1999;91(8):439–46.

21. Fisher JD, Fisher WA, Amico KR, Harman JJ. An information-motivation-behavioral skills model of adherence to antiretroviral therapy. Health Psychol. 2006;25(4):462–73.

22. Amico KR, Toro-Alfonso J, Fisher JD. An empirical test of the information, motivation and behavioral skills model of antiretroviral therapy adherence. AIDS Care. 2005;17(6):661–73.

23. Team L. The LifeWindows Information Motivation Behavioral Skills ART Adherence Questionnaire. 2006.

24. USAID. Measuring the Degree of HIV-related Stigma and Discrimination in Health Facilities and Providers. 2010.

25. Gombe MM, Cakouros BE, Ncube G, Zwangobani N, Mareke P, Mkwamba A, et al. Key barriers and enablers associated with uptake and continuation of oral pre-exposure prophylaxis (PrEP) in the public sector in Zimbabwe: Qualitative perspectives of general population clients at high risk for HIV. PloS one. 2020;15(1):e0227632–e.

26. Patel RC, Stanford-Moore G, Odoyo J, Pyra M, Wakhungu I, Anand K, et al. “Since both of us are using antiretrovirals, we have been supportive to each other”: facilitators and barriers of pre-exposure prophylaxis use in heterosexual HIV serodiscordant couples in Kisumu, Kenya. J Int AIDS Soc. 2016;19(1):21134.

27. Eakle R, Bothma R, Bourne A, Gumede S, Motsosi K, Rees H. “I am still negative”: Female sex workers’ perspectives on uptake and use of daily pre-exposure prophylaxis for HIV prevention in South Africa. PLOS ONE. 2019;14(4):e0212271.

28. Eakle R, Bourne A, Mbogua J, Mutanha N, Rees H. Exploring acceptability of oral PrEP prior to implementation among female sex workers in South Africa. Journal of the International AIDS Society. 2018;21(2):e25081.

29. Krishnaratne S, Bond V, Stangl A, Pliakas T, Mathema H, Lilleston P, et al. Stigma and Judgment Toward People Living with HIV and Key Population Groups Among Three Cadres of Health Workers in South Africa and Zambia: Analysis of Data from the HPTN 071 (PopART) Trial. AIDS Patient Care STDS. 2020;34(1):38–50.

30. Nyblade L, Addo NA, Atuahene K, Alsoufi N, Gyamera E, Jacinthe S, et al. Results from a difference-in-differences evaluation of health facility HIV and key population stigma-reduction interventions in Ghana. Journal of the International AIDS Society. 2020;23(4):e25483–e.

31. Duby Z, Fong-Jaen F, Nkosi B, Brown B, Scheibe A. ‘We must treat them like all the other people’: Evaluating the Integrated Key Populations Sensitivity Training Programme for Healthcare Workers in South Africa. South Afr J HIV Med. 2019;20(1):909-.

32. Mayer KH, Agwu A, Malebranche D. Barriers to the Wider Use of Pre-exposure Prophylaxis in the United States: A Narrative Review. Adv Ther. 2020;37(5):1778–811.

33. Opuni M, Bishai D, Gray GE, McIntyre JA, Martinson NA. Preferences for characteristics of antiretroviral therapy provision in Johannesburg, South Africa: results of a conjoint analysis. AIDS Behav. 2010;14(4):807–15.

34. Micheni M, Kombo BK, Secor A, Simoni JM, Operario D, van der Elst EM, et al. Health Provider Views on Improving Antiretroviral Therapy Adherence Among Men Who Have Sex with Men in Coastal Kenya. AIDS Patient Care STDS. 2017;31(3):113–21.

